# Predicting COVID-19 Pandemic in Saudi Arabia Using Modified Singular Spectrum Analysis

**DOI:** 10.1101/2020.05.24.20111872

**Authors:** Nader Alharbi

## Abstract

This research presents a modified Singular Spectrum Analysis (SSA) approach for the analysis of COVID-19 in Saudi Arabia. We have proposed this approach and developed it in [1–3] for separability and grouping step in SSA, which plays an important role for reconstruction and forecasting in the SSA. The modified SSA mainly enables us to identify the number of the interpretable components required for separability, signal extraction and noise reduction. The approach was examined using different number of simulated and real data with different structures and signal to noise ratio. In this study we examine its capability in analysing COVID-19 data. Then, we use Vector SSA for predicting new data points and the peak of this pandemic. The results shows that the approach can be used as a promising one in decomposing and forecasting the daily cases of COVID-19 in Saudi Arabia.

## 1 Introduction

One of the main issues that threats our health around the glop are infectious diseases. Nowadays, the outbreak of 2019 new coronavirus (SARS-CoV-2) disease (COVID-19) is a global pandemic [4, 5]. The first case of this virus was recognized and reported on 31-12-2019 in the city of Wuhan, the capital of Hubei in China [6]. Then, the virus has spread rapidly around the world and affected more than 200 countries [7].

The number of cases and deaths of this virus are globally considered as serious problems. The number of confirmed cases were more than 4 million and around 200 thousand deaths by 12-05-2020. Although the outbreak seems to have decreased in China, the virus and its impacts are still going global, and those numbers are increasing. This leads to our concerns about variation in the affected cases and the mortality rate of the COVID-19 pandemic. Furthermore, there are a lot of concerns about economic global impact of this crises. It is now understood that the devastating influence of the virus on economy and world health is incomparable [8].

The primary objective of this manuscript is construction of a reliable, robust and interpretable model describing, decomposing, forecasting the number of confirmed cases, and predict the peak of this pandemic in Saudi Arabia. The rate of mortality in Saudi Arabia is low, less than 1% till writing this paper. Thus, we are only interested in the new daily cases affected by the virus and try to detect its peak. The number of cumulative cases is more than 40000 by 12-05-2020.

There are many standard epidemiological models for modelling epidemics such SIR, see e.g. [9–11]. However, since our aim is to analyse the daily data series of COVID-19, we seek to use a promising, reliable, and capable method for analysing time series. There is a number of various methods for analysing time series, but several of these methods requiring, for example, linearity or non linearity of a particular form as they are parametric methods.

An alternative method uses non-parametric approaches that are neutral with respect to problematic areas of specification, such as linearity, stationarity and normality [12]. Thus, such approaches can show a reliable and better means of decomposing time series data. Singular Spectrum Analysis (SSA) is a relatively new non-parametric technique that has shown and proved its capabible use in several applications of time series in different disciplines, such as genetics and biology [13, 14], medicine [15, 16], engineering [17, 18], economics and finance [19, 20], and other areas. For its history, see [21,22]. For more details on the theory of SSA and its applications, refer to [23,24]. A comprehensive review of the method and description of its extensions and modifications can be found in [25].

Although the signals can be affected by an internal or external noise, which often have unknown characteristics, they can be identified if the signal and noise subspaces are accurately separated. It is known that removing noises from any signal is necessary for analysing any kind of time series, and is helpful in decomposing the signal in a proper manner [26].

The main idea of SSA is to analyse the main series into different components, then reconstruct the noise free series for further analysis. It depends upon two main choices; namely, the window length *L* and the number of required eigenvalues, denoted by r, for reconstruction. Thus, an appropriate selection of *L* and *r* leads to a perfect analysis and separability between the time series components. It was discussed in [27] that for a series of length *N*, selecting *L* = *N*/4 is common practice.It also should be mentioned that *L* should be large enough, but not larger than half of the series [23]. In [28], it was shown that for a series of length *N* and the optimal selection of the number of eigenvalues r for reconstructing the signal, the appropriate value of the window length is median {1,…,*N*}. Despite various attempts that have been applied, there is no universal rule for obtaining optimal selections of *L* and *r*.

We have proposed an approach in [2, 3] for the selection of the value of r for noise reduction, filtering, and signal extraction in SSA. This has also been applied to the distinction of noise from chaos in time series analysis [29], and for the correction of noise in gene expression data [30]. In [3], we have developed the approach and introduced new criteria to the discrimination between epileptic seizure and normal EEG signals, the filtering of the EEG signal segments, and elimination of the noise included in the signal. The approach is mainly used to identify the required number of eigenvalues/singular values corresponding to the signal component, which depends on the distribution of the eigenvalues of a scaled Hankel matrix. The correlation between eigenvalues, the coefficients of skewness, kurtosis and variation of the eigenvalues distributions were proposed and proved to be new criteria for the separability between signal and noise components as they can split the eigenvalues into two groups [2]. Different simulated and real signals were used considering different signal to noise ratio in [2, 3], and evaluated to show the ability of the approach in the selection of *r*.

The remainder of this paper is structured as follows: the following section gives a short description of the modified SSA approach and its algorithm. In Section 3, we show that this approach can decompose a synthetic data into two main distinct subspaces. Section 4 presents the implementation of the approach in decomposing and reconstructing COVID-19 daily cases series. The section also presents the prediction of COVID-19 in Saudi Arabia using Vector SSA for the extracted signal by the modified SSA. Section 5 draws the conclusion of this paper with some ideas for future work.

## 2 The Modified SSA method

### 2.1 Review

This section presents a short description of the modified SSA used in this manuscript (for more details refer to [2]). A time series is decomposed by the SSA technique into a sum of components, allowing the identification of each one as either a main or noise component. The goal here is to consider the signal as a whole so that we can identify the appropriate value of *r* related to the whole signal component. In other words, we are not interested in each signal component, so the selection of *L* rational to the periodicity of the signal components becomes less important [24]. Therefore, the modified SSA focuses on the selection of r to identify the signal subspace.

Consider a one-dimensional series *Y*_*N*_ = (*y*_1_,…, *y*_*N*_) of length *N*. Transferring this series into a multi-dimensional series *X*_1_, …, *X*_*K*_ where *X*_*i*_ = (*y*_*i*_, …, *y*_*i*+*L-*1_)^*T*^ ∈ **R**^*L*^ provides 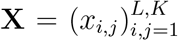, where *L* is an integer (2 ⩽ *L* ⩽ *N*/2) and *K* = *N - L* + 1. A matrix **X** is a Hankel matrix, all the elements along the diagonal *i* + *j* = *const* are equal. Set **B** = **XX**^*T*^ and denote by λ_*i*_ (*i* = 1, …, *L*) the eigenvalues of **B** taken in decreasing order of magnitude (λ_1_ ≥ … ≥ λ_*L*_ ≥ 0) and by *U*_1_, …, *U*_*L*_ the orthonormal system of the eigenvectors of matrix **B** corresponding to these eigenvalues.

The SVD of matrix **X** can be written as follows:

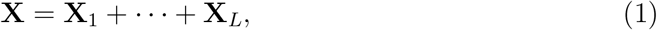

Where 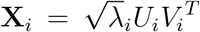. The elementary matrices **X**_*i*_ having rank 1, *U*_*i*_ and *V*_*i*_ are the left and right eigenvectors of matrix **X**. Note that the collection 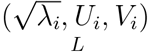 is called the ith eigentriple of the SVD. Note also that 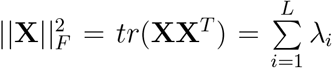 and 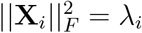,where || ||_*F*_ denotes the Frobenius norm.

Fundamental to the question of the eigenvalues behaviour, λ_*i*_, is that if the series size increases, there is a corresponding increase in the eigenvalues. This problem can be overcome if **B** is dividing by its trace, **A** = **B**/*tr*(**B**), which provides several important properties [1]. Let *ζ*_1_,…, *ζ*_*L*_ denote the matrix **A** eigenvalues in decreasing order of magnitude (1 ≥ *ζ*_1_ ≥ … ≥ *ζ*_*L*_ ≥ 0). The simulation technique is performed to obtain the distribution of *ζ*_*i*_ and to understand the behaviour of each eigenvalue. This helps to identify the value of r. Here, the goal is to establish the distribution and related forms of *ζ*_*i*_, which will be used to select the appropriate value of *r* for removing noise from COVID-19 series.

It was proved in our work [2] that the largest eigenvalue has a positive skewed distribution for a white noise process. Therefore, if skew(*ζ*_*c*_) (*c* ∈*{*1, …, *L}*) is the maximum, and the pattern for skew(*ζ*_*c*_) to skew(*ζ*_*L*_) has the same pattern, the same as emerged for the white noise, then the first *r* = *c −* 1 eigenvalues correspond to the signal and the rest to the noise. A similar procedure can be done using the the coefficients of kurtosis and variation of *ζ*_*i*_. Furthermore, if *ρ*_*s*_(*ζ*_*c−*1_, *ζ*_*c*_) is the minimum, and the pattern for the set 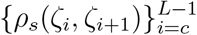 is similar to what was observed for the white noise, then we select the first r = c *−* 1 eigenvalues for the signal and the rest for the noise component (for more information see [2]).

In this research, we use the third and fourth central measures moments of the distribution, which are the skewness (*Skew*) and kurtosis (*Kurt*). Skewness is a measure of asymmetry of the data distribution, whilst kurtosis describes the distribution of observed data in terms of shape or peak. We use these measures as criteria for choosing the value of *r*, which can be calculated for *m* simulation as follows:

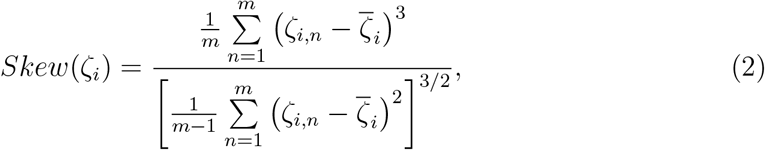

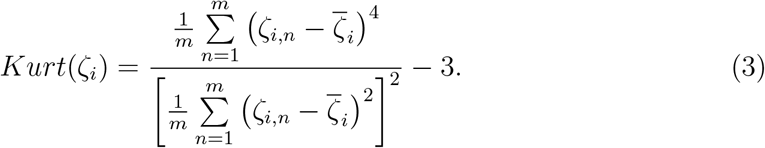

Moreover, the coefficient of variation, (*CV*), which is defined as the ratio of the standard deviation a(*ζ*_*i*_) and 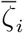 can be calculated mathematically from the following formula:

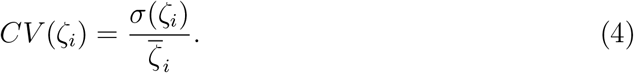

In addition, the Spearman correlation *ρ*_*s*_ between the eigenvalues *ζ*_*i*_ and *ζ*_*j*_ (*i, j* = 1,…, *L*) is also calculated to enhance the results obtained by those measures:

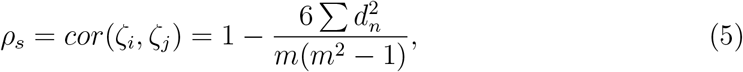

where *d*_*n*_ = *x*_*n*_ *− y*_*n*_ (*n* = 1,…, *m*) is the difference between x_*n*_ and y_*n*_ which are the ranks of *ζ*_*i,n*_ and *ζ*_*j,n*_ respectively, and *ζ*_*i,n*_ is the *n*-th observation for the *i*-th eigenvalue 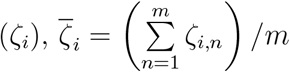.

These measures of difference between the eigenvalues related to the signal and noise components can specify the cut-off point of separability; the number of leading SVD components that are separated from the residual. Thus, the last cut-off point of separability between the signal and noise components obtained by the suggested measures, corresponds to the rank estimation.

The eigenvalues can be split into two groups by using the above criteria; the first corresponds to the signal and the second to the noise component. Furthermore, the Spearman correlation *ρ* between *ζ*_*i*_ and *ζ*_*j*_ is also calculated to support the outcomes obtained by those measures. The absolute value of the correlation coefficient is considered; 1 shows that *ζ*_*i*_ and *ζ*_*j*_ have perfect positive correlation, whilst 0 indicates there is no correlation between them. The matrix of the absolute values of the Spearman correlation gives a full analysis of the trajectory matrix, and in this analysis each eigenvalue corresponds to an elementary matrix of the SVD. Note that if the absolute value of *ρ* is close to zero, then the corresponding components are almost orthogonal; however, if it is close to one, then the two components are far from being orthogonal and so it is difficult to separate them. Thus, if *ρ* = 0 between two reconstructed components, this shows that these two reconstructed series are separable. The results of *ρ* between the eigenvalues for the white noise are quite large (see [2]), which helps in the discrimination of the noise part.

Once *r* is identified, then the matrices **X**_*i*_ can be split into two groups. Therefore, equation (1) can be written as follows:

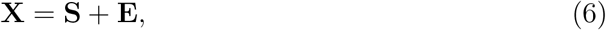

where 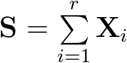 is the signal matrix and 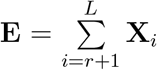 is the noise one. We then use diagonal averaging to transform matrix **S** into a new series of size *N* (see [23]).

### 2.2 Algorithm

The algorithm consists of two main stages. The steps of the first stage using the coefficients of skewness, kurtosis, variation and correlation can help us to obtain the optimal value of *r* for the separability between signal and noise as they split the eigenvalues into two groups. The steps of the second stage are used to reconstruct the free noise series

#### 2.2.1 Stage 1

1. Map a one-dimensional time series *Y*_*N*_ = (*y*_1_,…, *y*_*N*_) into multi-dimensional series *X*_1_,…, *X*_*K*_ with vectors *X*_*i*_ = (*y*_*i*_,…, *y*_*i*+*L-*1_)^*T*^ ∈ **R**^*L*^, where the window length *L* is an integer; 2 ≤ *L* ≤ *N*/2, and *K* = *N - L* + 1. This step gives us the Hankel matrix 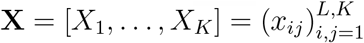.
2. Compute the matrix **A** = **XX**^*T*^ /*tr*(**XX**^*T*^).
3. Decompose matrix **A** as **A** = **PΓP**^*T*^, where **Γ** = diag(*ζ*_1_,…, *ζ*_*L*_) is the diagonal matrix of the eigenvalues of **A** that has the order (1 ≥ *ζ*_1_ ≥ *ζ*_2_,…, *ζ*_*L*_ ≥ 0) and **P** = (*P*_1_, *P*_2_,…, *P*_*L*_) is an orthogonal matrix whose columns are the corresponding eigenvectors.
4. Simulate the original series m times and calculate the eigenvalues for each series. We simulate y_*i*_ from a uniform distribution with boundaries *y*_*i*_ *−* a and *y*_*i*_ + *b*, where *a* =| *y*_*i−*1_ *− y*_*i*_ | and *b* =| *y*_*i*_ *− y*_*i*+1_ |.
5. Compute the skewness coefficient for each eigenvalue, *skew*(*ζ*_*i*_). If *skew*(*ζ*_*c*_) is the maximum, and the pattern for *skew*(*ζ*_*c*_) to *skew*(*ζ*_*L*_) has a similar pattern to the white noise, select *r* = *c −* 1.
6. Compute the coefficient of kurtosis for each eigenvalue, *kurt*(*ζ*_*i*_). If *kurt*(*ζ*_*c*_) is the maximum, select r = c *−* 1.
7. Compute the coefficient of variation, *CV* (*ζ*_*i*_). The result of *CV* splits the eigenvalues in two groups, from *ζ*_1_ to *ζ*_*c−*1_ which correspond to the signal, and the remainder, which have an almost U shape, correspond to the noise.
8. Compute the absolute values of the correlation matrix between the eigenvalues, and represent them in a 20-grade grey scale from white to black corresponding to the values of the correlations from 0 to 1. This matrix also splits the eigenvalues into two groups, from *ζ*_1_ to *ζ*_*r*_ which correspond to the signal, and the remainder, which correspond to the noise.

#### 2.2.2 Stage 2

1. Calculate the approximated signal matrix 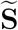, that is 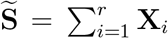, where *r* is obtained from the first stage, 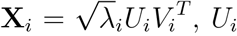 and *V*_*i*_ stands for the left and right eigenvectors of the trajectory matrix.
2. By averaging over the diagonals of matrix 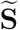, this gives a one dimensional series, which is the approximate signal 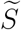.

The capability of the modified SSA using different synthetic data, including series generated from chaotic map systems with different Signal to Noise ratio (SNR), were presented in [2]. This result confirms that the approach works promisingly for any series that is mixed with a low or high noise level.

Each eigenvalue or singular value contributes to the trajectory matrix decomposition. We cansider the ratio 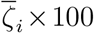 to be the characteristic of matrix **H**_*i*_ to Eq. (1). Thus, 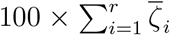 is considered as the characteristic of the optimal approximation of **H** by matrices of rank *r*.

## 3 Separability in Synthetic data

We should mention that using the standard criteria in the basic SSA, the weighted correlation or *w* -correlation for separability and grouping (for more information see [24]), does not always provide a good separability and correct selection of *r*, specially for real data.

It was shown in [2] that the results based on *Skew, Kurt, CV*, and *ρ*_*s*_ are more accurate than those obtained by the *w* -correlations for small window length, particularly for a data where a linear trend is included in the series.

Thus, we use the modified SSA, in particular, we use some of those proved criteria on the distribution of *ζ*_*i*_, which given in the previous sections to identify *r*. The results are plausible and reliable.

We will provide here one synthetic example to show the capability of the approach before applying it to COVID-19 data, for more examples considering different types of series and evaluations with different criteria refer to [2].

### Simulated data

In the following example, a white noise process *ϵ*_*t*_ was added to an exponential trend series.

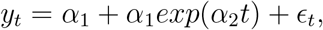

where *t* = (1,…, *N*), *N* = 42, *ʱ*_1_ = 10, *ʱ*_2_ = 0.09, and *ϵ*_*t*_ is a Gaussian white noise process with variance 1 (see 1). It is obvious that the number of eigenvalues required to reconstruct the signal for this series is 2, as we have a constant adding to exponential curve, which corresponds to the rank estimation (see [23].

By looking at the *w* -correlations, and the logarithm of the eigenvalues, we may use only the first component to extract the signal (see Fig. 2).

**Figure 1:**
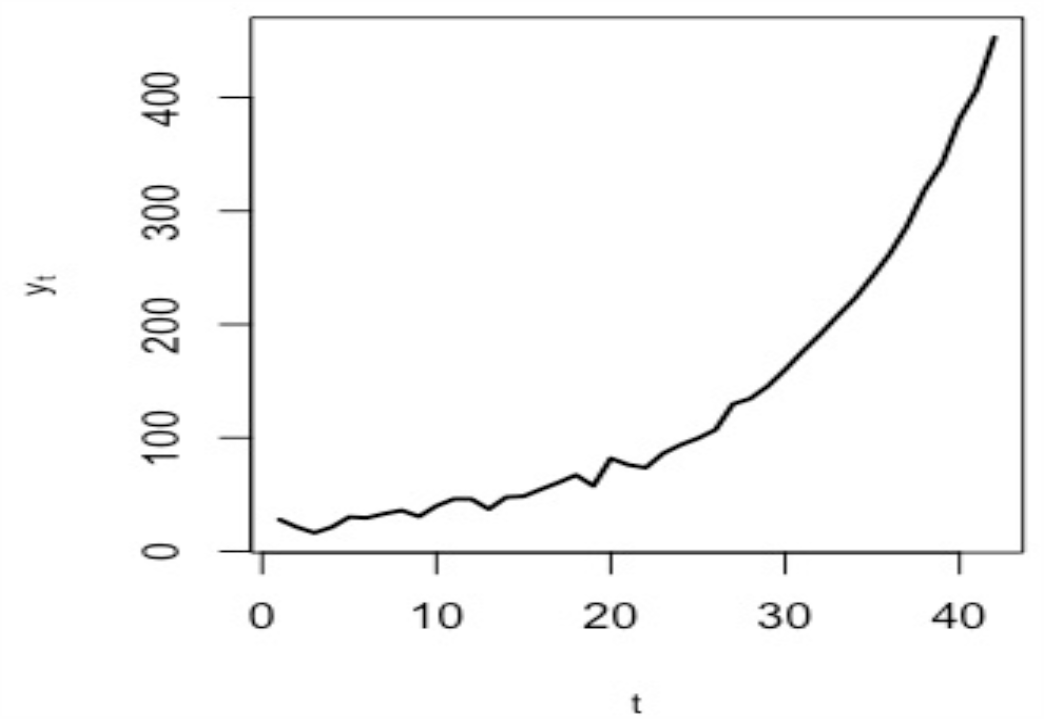
A realization of the simulated series.

**Figure 2:**
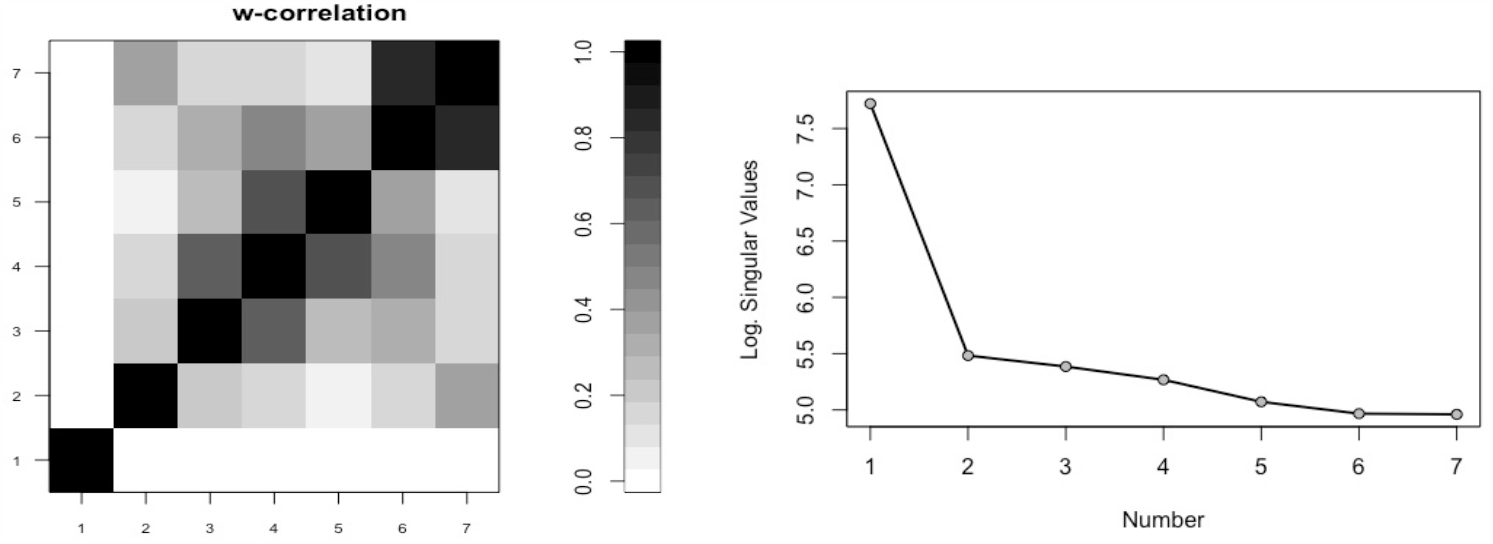
w-correlations matrix (left) for the 7 reconstructed components of the simulated series, and logarithms of the 7 simulated series eigenvalues (right).

However, using the suggested measures and criteria, this gives us the correct value of *r*. Fig. 3 represents the kurtosis coefficient of *ζ*_*i*_ (*i* = 1,…, *L*). The maximum value of the kurtosis coefficient is considered as one of the rules and indicators we use for the start of the noise. It is clear that the maximum kurtosis coefficients of *ζ*_*i*_ is obtained for *ζ*_*c*=3_. Thus, the number of eigenvalues required to extract the signal is *r* = *c −* 1 = 2. Similar results emerged by using the values of *skew* and *CV* (see Fig 4).

**Figure 3:**
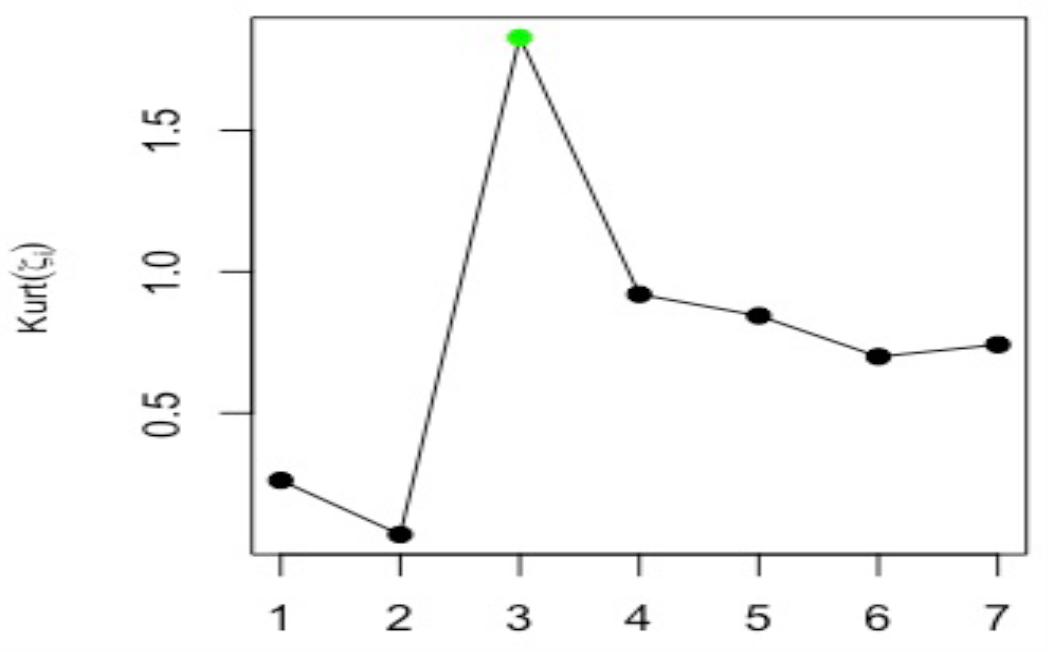
Kurtosis of *ζ*_*i*_ for the simulated series.

**Figure 4:**
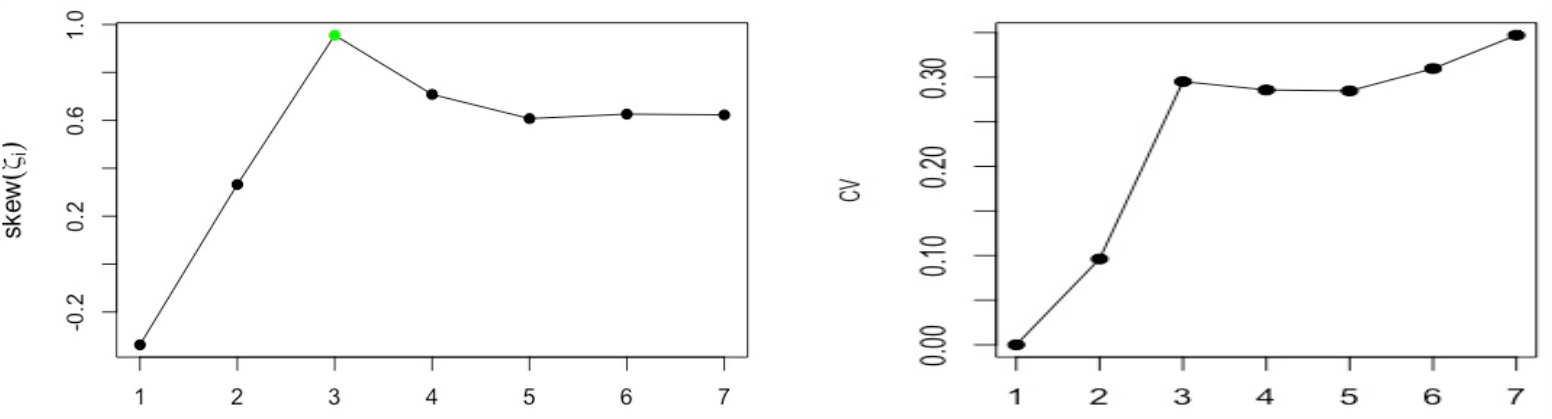
Skewness (left) and coefficient of variation (right) of *ζ*_*i*_ for the simulated series.

In addition, the Spearman correlation coefficient between *ζ*_*i*_ and *ζ*_*i*+1_ is also calculated. Fig. 5 (left) shows the correlation between *ζ*_*i*_ and *ζ*_*i*+1_. For the correlation coefficient, the minimum value of *ρ*_*s*_ between *ζ*_*c−*1_ and *ζ*_*c*_ is used as an another indicator for the cut-off point. The results are similar to what emerged by other criteria, and confirm that the approach works properly. Different criteria were used in [2] to evaluate the approach, for example, RMSE and MAE, which confirm that the modified approach can be used as a promising one.

**Figure 5:**
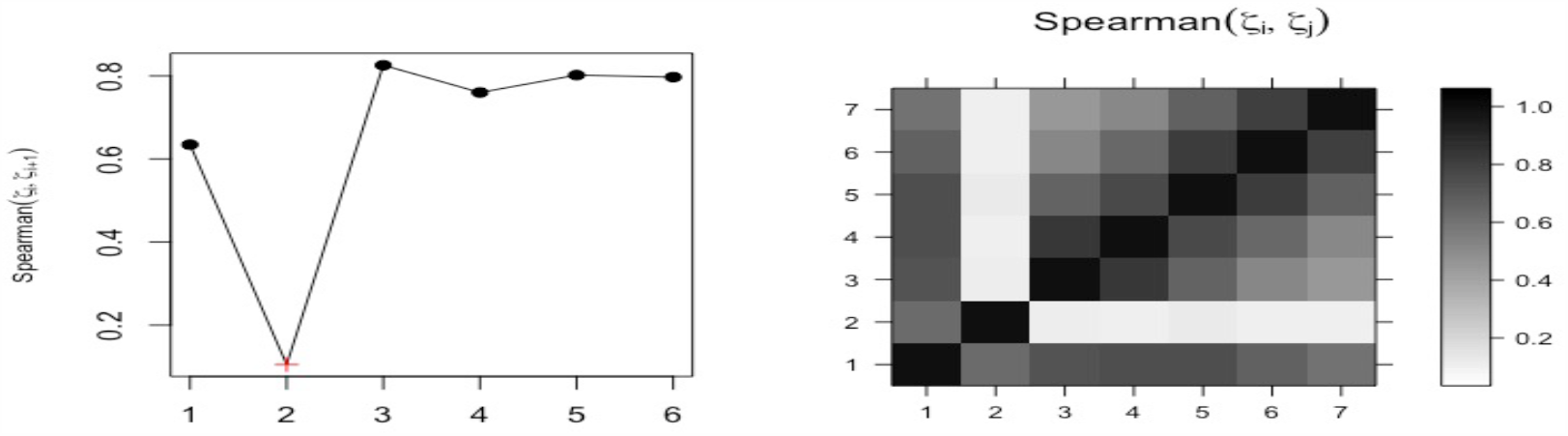
Spearman correlation of (*ζ*_*i*_, *ζ*_*i*+1_) (left) and matrix of Spearman correlation between (*ζ*_*i*_, *ζ*_*j*_).

The correlation matrix also enables us to distinguish and separate the different components from each other. Thus, the correlation matrix of *ζ*_*i*_ is identify the separability between the components. If the absolute value of the correlation coefficient between *ζ*_*i*_ and *ζ*_*j*_ is small, then the corresponding components are almost orthogonal; however, if the value is large, then the corresponding series are far from being orthogonal and thus they are not neatly separable. It is clear that the signal can be separated from the noise since the top right pattern from the correlation matrix is related to the white noise process (see Fig. 5 (right)).

## 4 COVID-19 data analysis

The daily confirmed cases of COVID-19 in Saudi Arabia [31] is used in this research. First, We have used the first 42 days data; from 02-03-2020 to 12-04-2020. The aim is to analyse the data and make prediction from 13-04-2020, and detect the peak. The number of daily cases series is depicted in Fig. 6. Second, we have updated our data on 20-05-2020, and included values from 13-04-2020 to 12-05-2020, so the total became 81 values. This does not affect the required number of eigenvalues for the reconstruction stage, this will be discussed in the following part.

**Figure 6:**
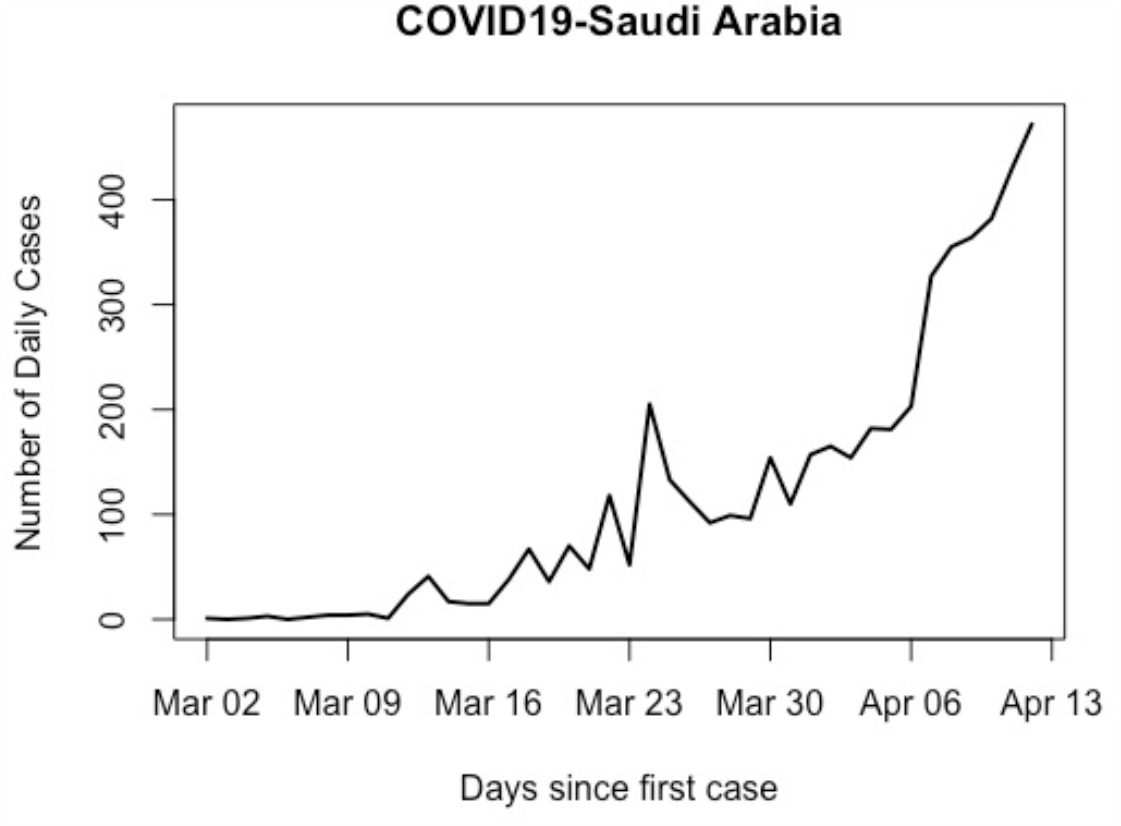
COVID-19 daily confirmed cases time series in Saudi Arabia (02-03-2020 to 12-04-20202)

### 4.1 Separability and selection of the components

let us now start with the first data. As we mentioned earlier, since our aim is to extract the signal as a whole, we can choose any value for *L*, and the goal to find the best choice of *r*. Furthermore, based on our research [2], we showed that one can use a small window length when analysing exponential series, like the one of COVID-19 series. The selection of *L* = 7, provide the best and reasonable results with the required *r* that will be obtained by the proposed approach.

Th results based on those measure in extraction the signal for forecasting, give a curve with likely peak. However, the prediction using many other choices of *L* and *r* do not give any end or peak for this pandemic and go up exponentially, and this is impossible as this pandemic will not stay forever. This also support the obtained results. Therefore, the important task now is the selection of the number of eigenvalues *r* that required for reconstruction and build the model for forecasting.

Fig. 7 illustrates the results of the coefficients of skewness and kurtosis for each eigenvalue, and the results of the matrix correlations and correlation between *ζ*_*i*_ and *ζ*_*i*+1_ for *L* = 7. As shown by the results, for the COVID-19 daily series, the maximum values of *Skew, Kurt*, are observed for *ζ*_*c*=3_, and the minimum value of *ρ*_*s*_ is obtained between *ζ*_*c−*1=2_ and *ζ*_*c*=3_. In addition, the matrix of Spearman correlation for *ζ*_*i*_ and *ζ*_*j*_ (*i, j* = 1,…, 7) split the eigenvalues or the components into two groups; this indicates that the value of *r* = 2.

**Figure 7:**
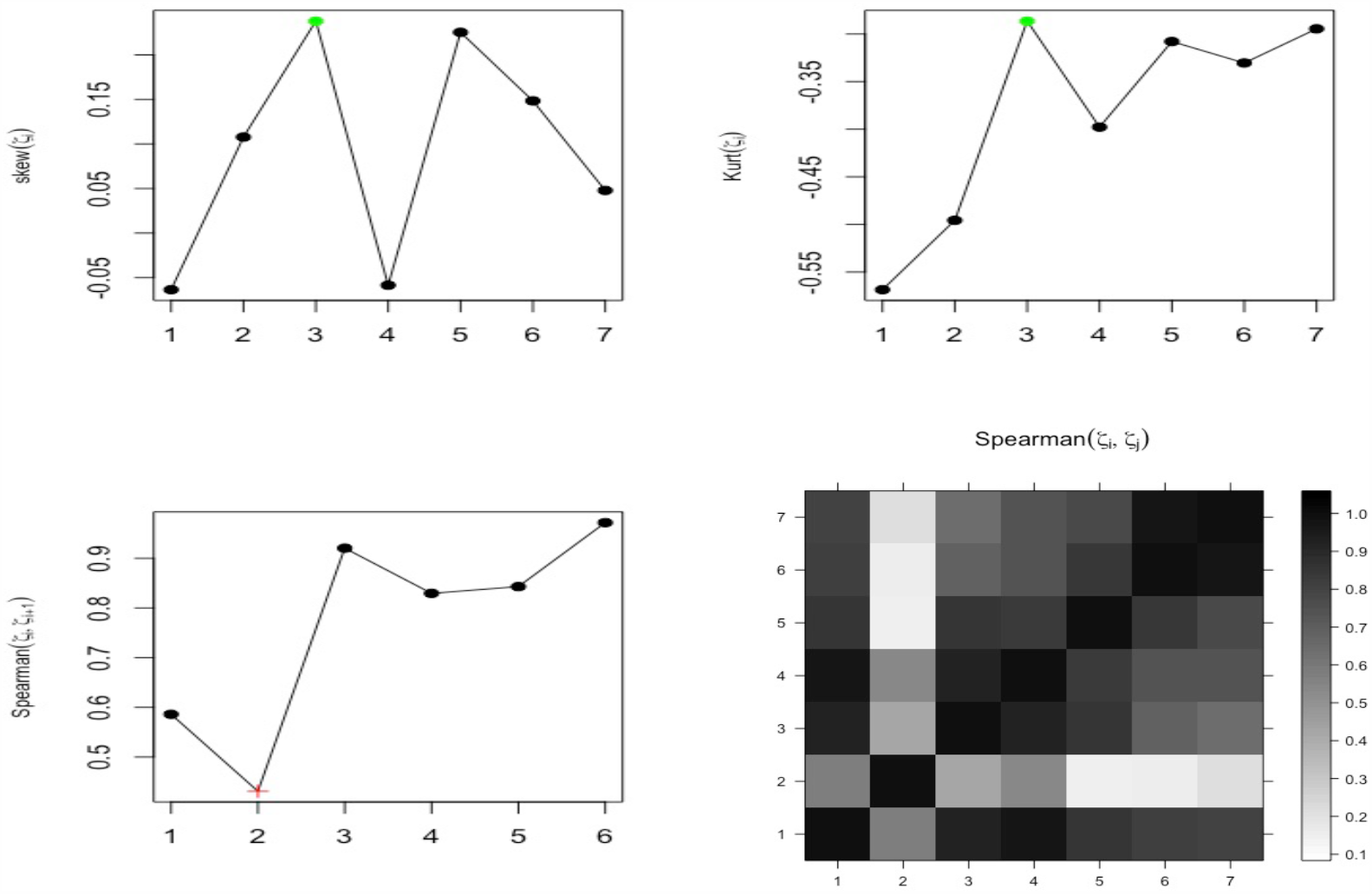
All measures results for *ζ*_*i*_.

Fig. 8 depicts the the result of the reconstructed series, which is obtained by using *L* = 7 and eigentriples *r* = 2. The red and the black lines correspond to the reconstructed series and the original series, respectively. It seems that the reconstructed series has been obtained well. However, we will see later that the the reconstructed series using the whole data is better than this fitted series.

**Figure 8:**
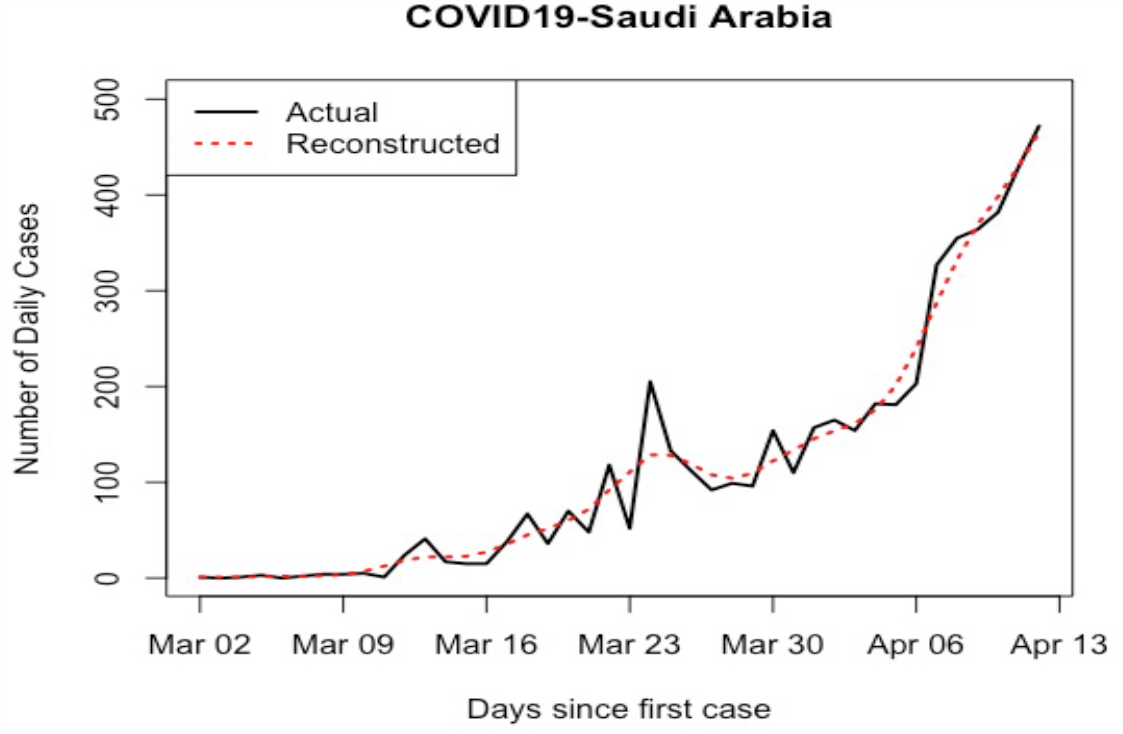
Plot of the daily Covid-19 series in Saudi Arabia and fitted curve.

### 4.2 Prediction daily cases of COVID-19 using VSSA

After obtaining the reconstructed series, the next aim is to predict new data, we will predict daily cases from 13-04-2020 to August 2020. There are two main forecasting methods in SSA, Vector SSA (VSSA) and Recurrent SSA (RSSA). The VSSA forecasting algorithms is the most widely used in SSA [23]. Generally, this method works more robustly than RSSA especially when a series contains outliers or when faced big shocks in the series [32]. Therefore, we use the VSSA algorithm for forecasting in this research as recommended in [12].

#### Vector forecasting algorithm

For performing SSA forecasting, the basic requirement is that the series satisfies a linear recurrent formula (LRF). The series *Y*_*N*_ = [*y*_1_,…, *y*_*N*_] satisfies a LRF of order *L −* 1 if:

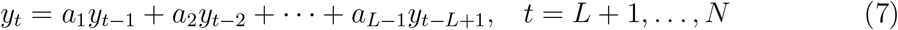

The coefficient vector *A* = a_1_,…, a_*L−*1_ is defined as follows:

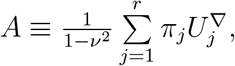

where 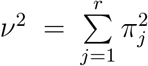 and 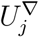 is the vector of the first *L −* 1 components of the eigen-vector *U*_*j*_, and *π*_*j*_ the last component of *U*_*j*_ (*j* = 1,…, *r*).

Consider the following matrix

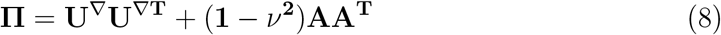

let us now define the linear operator:

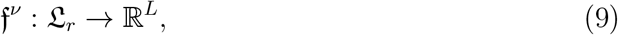

where 𝔏_*r*_ = span*{U*_1_,…, *U*_*r*_*}* and

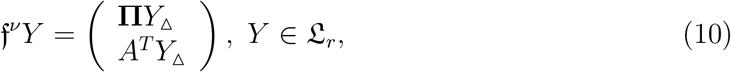

where *Y*_Δ_ is the vector of the last *L −* 1 elements of *Y*_*N*_. The vector *Z*_*j*_ is defined as follows:

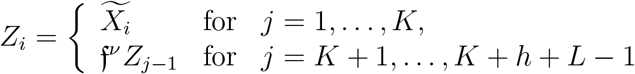

where the 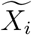 are the reconstructed columns of the trajectory matrix of the *i* th series after grouping and leaving out noise components. Now, by constructing matrix Z = [*Z*_1_,…, *Z*_*K*+*h*+*L−*1_] and performing diagonal averaging, a new series *ŷ*_1_,…, *ŷ*_*K*+*h*+*L−*1_ is obtained, where *ŷ*_*N*+1_,…,*ŷ*_*N*+*h*_ from the h terms of the VSSA forecast.

As we discussed above, the best values for reconstruction and forecasting are *L* = 7 and *r* = 2. Similar procedures have been done for the new data that updated on 20-05-2020. Same values of *L* and *r* were used in analysing the new data, and used for prediction new data points. Fig. 9 presents the updated data and the reconstructed series by the first 2 eigentriples. It is obvious that the reconstructed series is obtained precisely. Fig. 10 shows two curves predictions and the whole actual data, the red one is the prediction using the first data, and the blue one is the predictions using the updated data. It is obvious that there is no big difference, as the peak by the red curve around end of May where nearly end of June by the blue curve that used the updated data. In addition, the end of this pandemic can be between July and mid of August.

**Figure 9:**
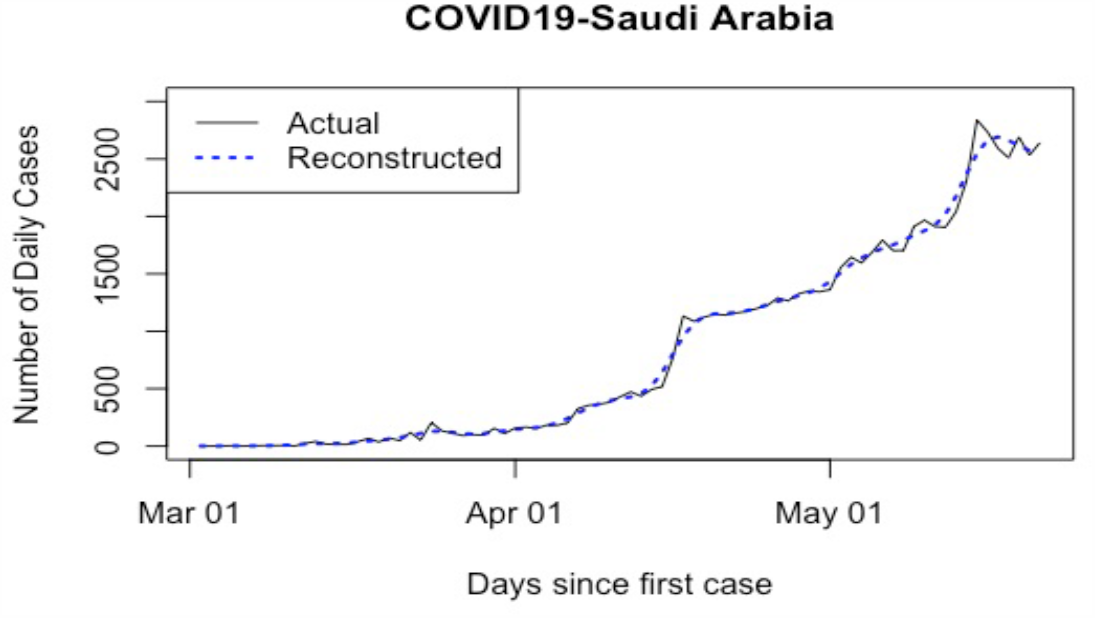
Plot of the daily COVID-19 series in Saudi Arabia and fitted curve for the whole data.

**Figure 10:**
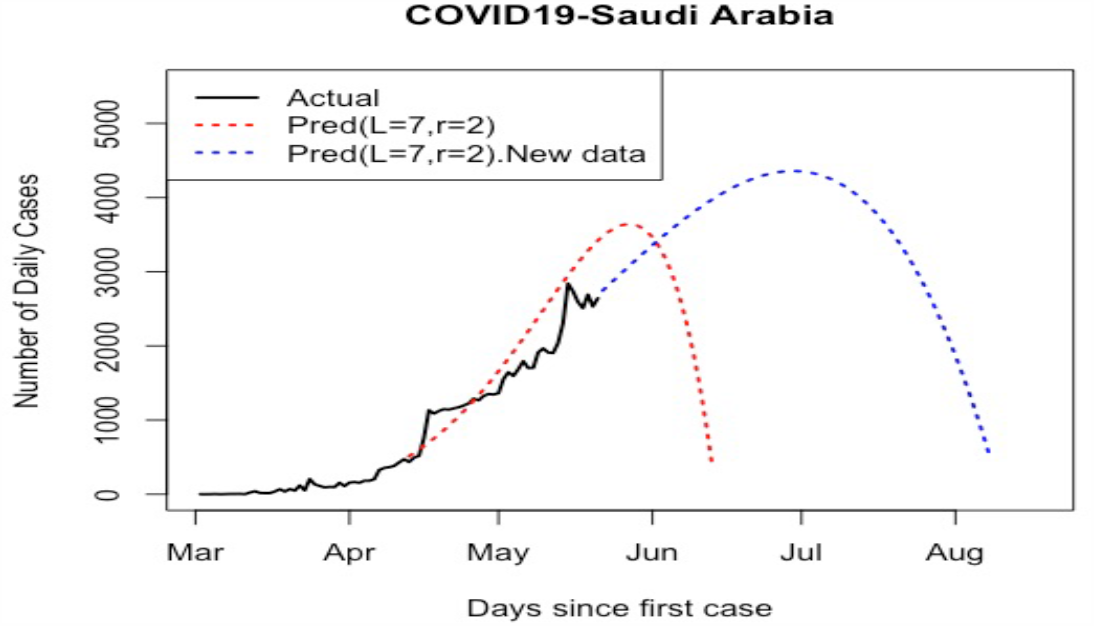
Comparison of two forecasting scenarios with actual observations.

## 5 Conclusion

A modified Singular Spectrum Analysis approach were used in this research for the decomposing and forecasting COVID-19 data in Saudi Arabia. The approach was examined in our previous research, and here in analysing COVID-19 data.

In the first stage, the first 42 confirmed daily values (02-03 to 12-04-2020) were used and analysed to identify the value of *r* for separability between noise and the signal. After obtaining the value of *r*, which was 2, and extracting the signals, the Vector SSA were used for prediction and determine the pandemic peak. In the second stage, we updated the data and included the 81 daily values. We have used the same window length and number of eigenvalues for reconstruction and forecasting. The results of both forecasting scenarios have indicated that the peak will be around end of May or end of June, and the end of this crises can be between end of June and mid of August.

All our results confirm the impressive performance of the modified SSA in analysing COVID-19 data and selecting the value of *r* for identifying the signal subspace from a noisy time series, and then make a good prediction using Vector SSA method. Note that we have not examined all possible values of window length in this research, and for forecasting we have used only the basic Vector SSA.

For future research, we will include more data and considered different window length *L* that may give a better forecasting. In addition, chaotic behaviour in COVID-19 data will be examined as we have some results that show strange patterns, which can be found in chaotic systems.

## Data Availability

The daily confirmed cases of COVID-19 in Saudi Arabia [32] is used in this research. First, We have used the first 42 days data; from 02-03-2020 to 12-04-2020.
https://covid19.moh.gov.sa

